# Continuous Time-Domain Near-Infrared Spectroscopy During Endovascular Thrombectomy Enables Early Prediction of Post-Recanalization Cortical Infarct

**DOI:** 10.64898/2026.04.20.26345790

**Authors:** Masih A. Rafi, Tigran Khachatryan, Shreen A. Abdel-Hadi, Sophia V. Hason, Amber N. Huynh, Janaka C. Ranasinghesagara, Rhaviel A. Rivera, Elina Khachatryan, Audrey N. Zhang, Shirin M. Dara, Jordan C. Xu, Kiarash Golshani, Frank P. Hsu, Shuichi Suzuki, Ichiro Yuki, Jefferson W. Chen, Imad R. Khan, Sara J. Stern-Nezer, Patrick M. Chen, Kenji Yoshimoto, Hiroko Wada, Etsuko Ohmae, Yukio Ueda, Wengui Yu, Vasan Venugopalan, Yama Akbari

## Abstract

**Background:** Existing neuromonitoring tools lack the capability for continuous, intraoperative assessment of cerebral oxygenation and tissue viability during endovascular thrombectomy (EVT) of large vessel occlusion stroke. Time-domain near-infrared spectroscopy (TD-NIRS), an advanced non-invasive optical technique, may overcome this challenge. However, no study has yet investigated TD-NIRS’ feasibility as an intraoperative neuromonitoring tool during EVT and its capability in predicting post-recanalization infarct.

**Methods:** In this prospective observational study, eleven patients with middle cerebral artery or internal carotid artery occlusion were monitored during EVT using TD-NIRS. Absolute concentrations of oxyhemoglobin ([HbO_2_]), deoxyhemoglobin ([HHb]), total hemoglobin ([tHb]), and tissue oxygen saturation (StO_2_) on both hemispheres was derived using a scaled Monte Carlo approach. Patients were dichotomized into post-EVT cortical infarct and no infarct, and the Δ[HbO_2_]/[HHb] pre- vs post-recanalization was assessed.

**Results:** Significant differences were observed in pre- vs post-recanalization [HbO_2_] (*p*=0.0068), [HHb] (*p*=0.042), and StO_2_ (*p*=0.0020) on the affected hemisphere. The Δ[HbO_2_]/[HHb] significantly differed between patients with and without post-EVT infarct (*p*=0.0043). Logistic regression (*p*=0.0041) and ROC (AUC=0.82) determined that Δ[HbO_2_]/[HHb] reliably predicts post-EVT cortical infarct.

**Conclusions:** These results suggest TD-NIRS as a novel, adjunctive intraoperative neuromonitoring tool during EVT, with potential to predict post-EVT cortical infarct, guiding clinical management before/after thrombectomy.

**Graphical Abstract:** 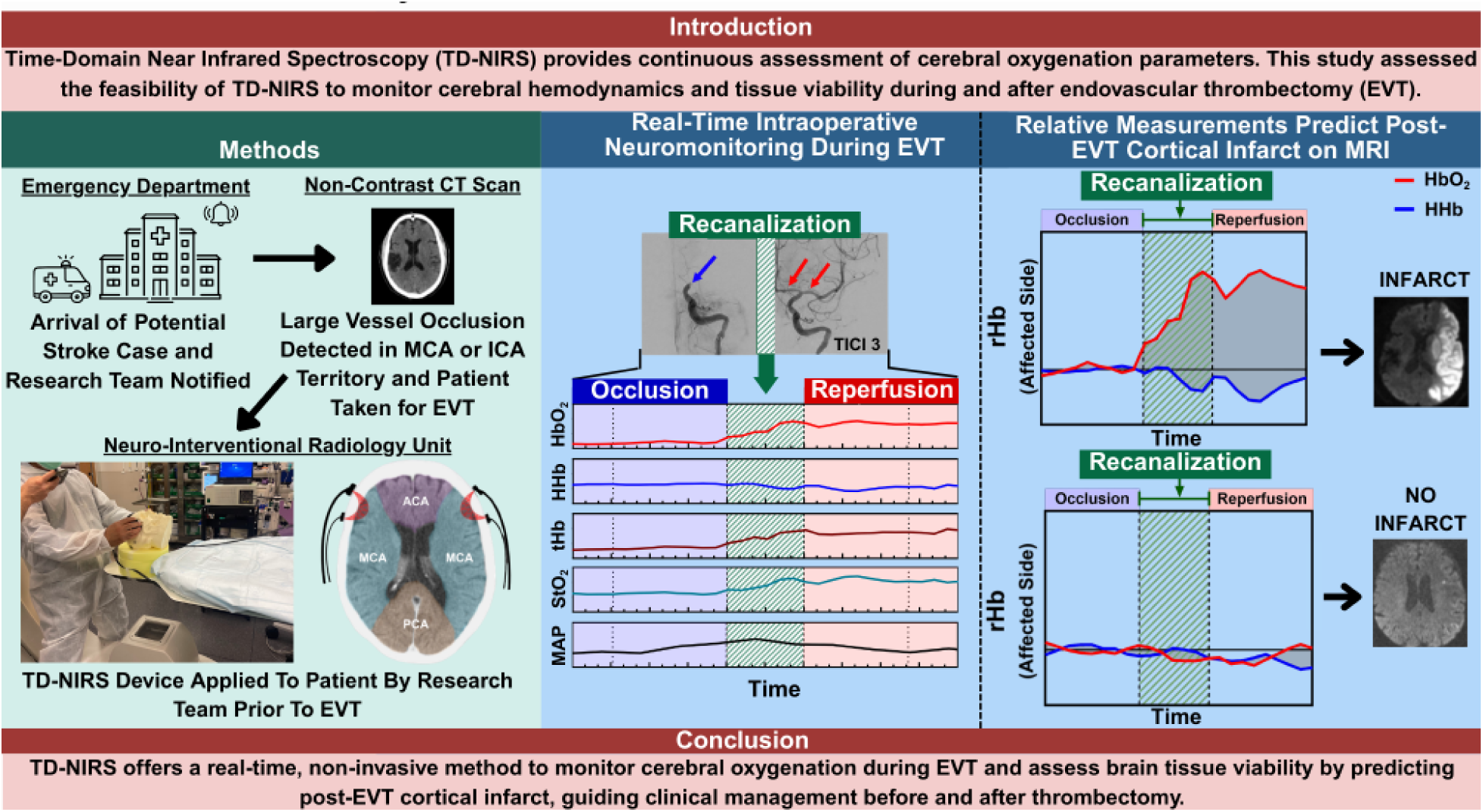

## Introduction

Acute ischemic stroke is a leading cause of morbidity and mortality worldwide. In 2019, the Global Burden of Disease, Injuries, and Risk Factors Study reported that stroke is the second leading cause of death worldwide, accounting for 11.6% of total deaths.(1) Approximately 24-46% of acute ischemic strokes are due to large vessel occlusion (LVO).(2) Although a substantial number of patients with LVO meet the eligibility criteria for endovascular thrombectomy (EVT), only 5-14% receive the procedure due to limited access, resource availability, and system-level constraints.(3,4) In 2015, five randomized trials showed that EVT is of significant outcome benefit to most patients with acute ischemic stroke caused by occlusion of the proximal anterior circulation, irrespective of patient characteristics or geographical location.(5)

Current diagnostic tools are limited in their capacity to provide reliable, continuous monitoring. Neurological evaluations, such as the National Institutes of Health Stroke Scale (NIHSS), assess stroke severity but are subjective and may be influenced by age, pre-existing comorbidities, and prior neurological conditions. Imaging modalities, including non-contrast computed tomography (CT) with the Alberta Stroke Program Early CT Score (ASPECTS), CT angiography (CTA), CT perfusion (CTP), and magnetic resonance imaging (MRI), are considered the standard of care for evaluating vascular occlusion and infarct burden. However, these modalities offer static snapshots of the brain’s perfusion status and cannot provide real-time, continuous neuromonitoring.(6–9) While vital signs—such as blood pressure (BP), heart rate (HR), peripheral oxygen saturation (SpO_2_), and electrocardiography—are utilized for comprehensive patient evaluation, they may not reflect cerebral hemodynamics due to impaired brain autoregulation and are of limited utility.(10,11) Other non-invasive techniques, such as electroencephalography, transcranial Doppler, somatosensory evoked potentials, and motor evoked potentials, can provide indirect measurements of cerebral perfusion. However, their results are often confounded by sedation, movement artifacts, anatomical variation, operator error, previous history of brain or spinal cord injury, and body temperature.(12–16)

While fluoroscopy remains the gold standard neuroimaging modality during EVT, it does not provide continuous assessment of microvascular blood flow and local oxygenation metrics.(17) Similarly, the Thrombolysis in Cerebral Infarction (TICI) grading system, although employed as standard of care, does not provide a dependable evaluation of end-organ reperfusion status or establish reliable prognostic value.(18–21) While continuous wave near-infrared spectroscopy (CW-NIRS) is increasingly used to provide relative measures of cerebral hemodynamics in clinical settings, no reliable technology has been established to continuously monitor cerebral perfusion and metabolism during EVT.(22,23) Given that both ischemic and reperfusion phases are critical in patients with acute ischemic stroke, these limitations underscore the need for an adjunct, non-invasive neuromonitoring tool for continuous evaluation of cerebral tissue perfusion and metabolism throughout EVT.(24)

Time-domain near-infrared spectroscopy (TD-NIRS) is an advanced optical approach that measures the temporal dispersion of short pulses of light as they travel from an optical source to a receiver. This measurement of temporal dispersion allows for the extraction of absolute values for tissue optical properties parameterized by tissue absorption (*μ*_a_) and reduced scattering (*μ*′_s_) coefficients. Once these properties are determined at multiple wavelengths, absolute values of chromophore concentrations within the probed tissue volume (oxyhemoglobin [HbO_2_] and deoxyhemoglobin [HHb]) can be computed.(25,26) Several research groups have demonstrated the utility of TD-NIRS as a neuromonitoring tool by measuring absolute values of cerebral perfusion and oxygen metabolism in acute stroke patients using laboratory or commercial TD-NIRS systems at bedside. A 2012 study measured the inter-hemispheric perfusion difference in 10 patients with acute stroke using a dynamic contrast-enhanced TD-NIRS system.(27) Another study employed TD-NIRS to measure [HbO_2_], [HHb], [tHb], and tissue oxygen saturation (StO_2_) in 47 patients with acute ischemic stroke within 24 hours, and again after 24 hours of stroke onset, which found pronounced differences in [HHb], [tHb], and StO_2_ between patients and controls.(28) Yet another study used Hamamatsu’s TD-NIRS (TRS-20) system to measure [HbO_2_], [HHb], [tHb], StO_2_, in 5 patients with chronic stroke and found decreases in [HHb] and increases in StO_2_ on the affected side, indicating reduced O_2_ consumption.(29) Despite these successes, no studies have investigated TD-NIRS for use as an adjunct *<u>intraoperative</u>* neuromonitoring tool during EVT.

In the present study, we aimed to evaluate the feasibility of intraoperative TD-NIRS monitoring of patients with acute stroke undergoing EVT. We used Hamamatsu’s TRS-21 system to measure TD-NIRS signals and employed a custom-developed Monte Carlo algorithm to recover cerebral hemoglobin metrics ([HbO_2_], [HHb], [tHb]) and StO_2_.(30) Unlike conventional methods, our approach utilizes a transport-rigorous scaled Monte Carlo (sMC) approach to provide improved accuracy in estimating cerebral optical properties from TD-NIRS signals during EVT.(31,32)

## Materials and Methods

### Study Design

This prospective observational cohort study was conducted to assess the feasibility of TD-NIRS as a continuous, non-invasive neuromonitoring system in patients with acute ischemic stroke undergoing EVT. The study was approved by the Institutional Review Board (IRB) at the University of California, Irvine (IRB#20195366), and written informed consent was obtained from the legally authorized representatives of all patients. This study was conducted in accordance with the Declaration of Helsinki, the Health Insurance Portability and Accountability Act (HIPAA), and Good Clinical Practice (GCP) principles. Data acquisition was standardized, raw TD-NIRS signals were screened for artifacts prior to analysis, and the data analysis team was blinded to clinical outcomes. Results were reported in accordance with Strengthening the Reporting of Observational Studies in Epidemiology (STROBE) guidelines.

### Inclusion/Exclusion Criteria

The stroke team adhered to the 2019 American Heart Association (AHA) guidelines for selecting patients for EVT. These guidelines emphasize rapid and accurate identification of patients with emergent LVO within the anterior circulation who can benefit from EVT.(33) Patients eligible for inclusion in this study were ages 18 years or older, diagnosed with acute ischemic stroke due to LVO in the middle cerebral artery (MCA) or internal carotid artery (ICA), underwent EVT, achieved recanalization scores ranging from TICI 2b (near-complete reperfusion) to TICI 3 (complete reperfusion with filling of all distal branches). The measured patients presented within 6-24 hours of symptom onset and were selected for EVT based on clinical and neuroimaging assessments. Non-contrast CT was used to exclude intracranial hemorrhage and evaluate infarct extent (e.g., favorable ASPECTS), while CTP, when performed, was used to estimate infarct core and identify potential salvageable brain tissue. Informed consent from patient or legally authorized representative (LAR) prior to EVT was required.

### Timeline of Events

The timeline for each patient (Figure 1) began with the arrival of first responders to the stroke scene, where a stroke assessment, based on the Los Angeles Motor Scale (LAMS) score, was conducted. Our clinical research team was notified regarding the potential stroke case when the hospital was alerted about the patient’s arrival and/or when the patient arrived at the emergency department and underwent a comprehensive stroke evaluation. Standard protocol for acute stroke workup was subsequently followed, including non-contrast head CT and decision-making for intravenous thrombolysis therapy with tenecteplase (TNK) and EVT. For patients who met the inclusion criteria and underwent EVT, TD-NIRS monitoring was implemented in the neuro-interventional radiology unit. Mechanical thrombectomy was performed utilizing A Direct Aspiration First Pass Technique (ADAPT) except for tandem occlusions. A stent retriever was used, when necessary, based on the neurointerventional radiologist’s decision. For tandem occlusions, preference was given to stent-retriever thrombectomy with additional balloon angioplasty of cervical ICA occlusion according to established institutional protocol. TICI scores were used to evaluate the extent of reperfusion achieved, with scores assessed by the neuro-interventional radiology team to determine procedural success. TICI score was measured based on degree of perfusion in downstream territory of the occluded vessel, with scores ranging from 0 (no perfusion) to 3. Following recanalization, continuous TD-NIRS monitoring was performed to assess post-procedural cerebral oxygenation and hemodynamics at bedside in the intensive care unit.

**Figure 1.**
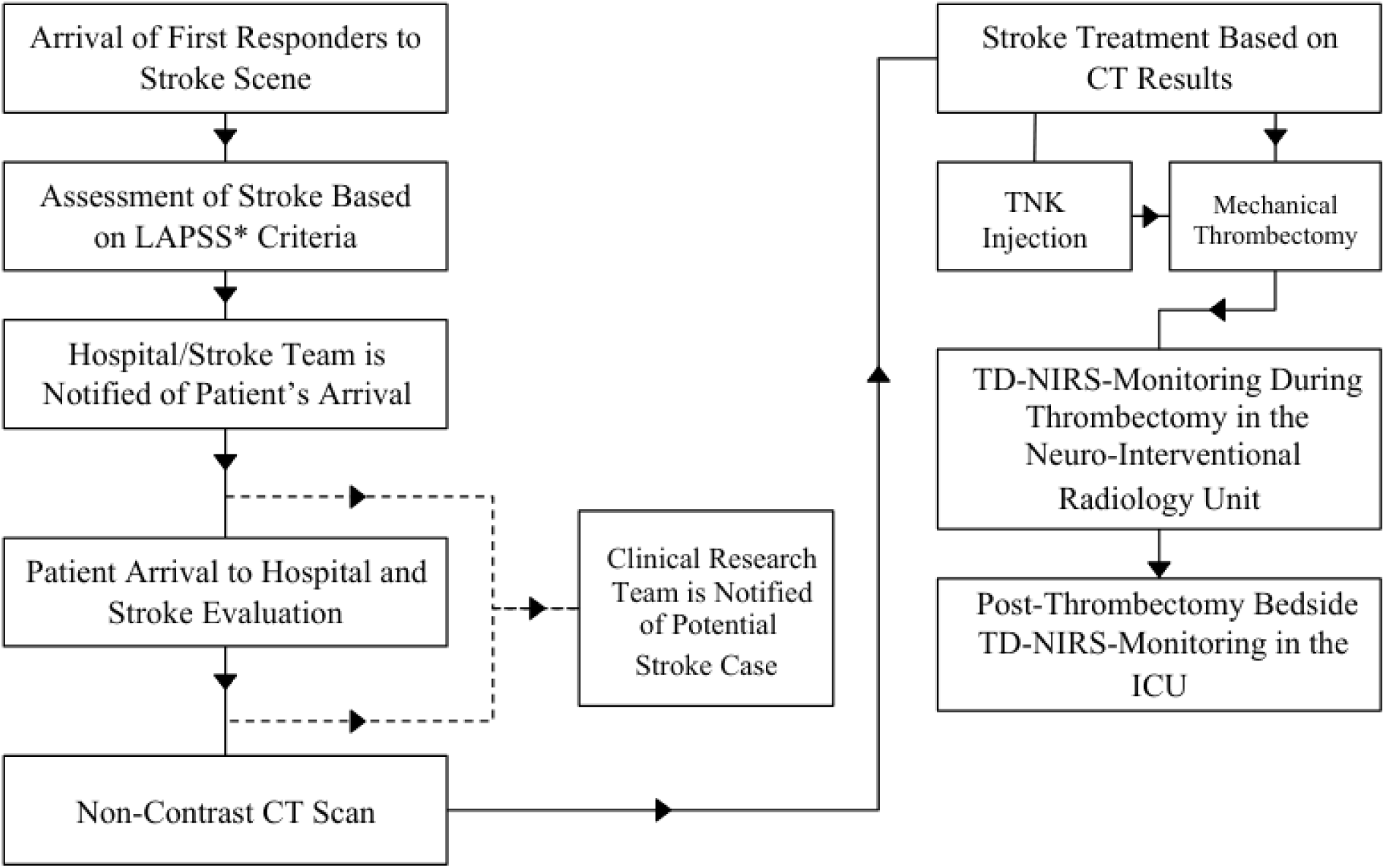
Schematic representation of the study workflow. Abbreviations: LAPSS, Los Angeles Prehospital Stroke Screen; CT, computed tomography; TNK, tenecteplase; TD-NIRS, Time-Domain Near-Infrared Spectroscopy; ICU, intensive care unit.

### Data Collection and Processing

Our TD-NIRS system (Hamamatsu’s TRS-21) provides three wavelengths (760, 800, 830 nm) of pulsed near-infrared light and measures the time-correlated diffusion of photons from the optical source to the detector. Using this data, we derive *μ*_a_ and *μ*′_s_ coefficients using a sMC approach.^32^ Absolute hemoglobin concentrations ([HbO_2_], [HHb], [tHb]), and StO_2_ are then determined from these coefficients. Upon meeting the inclusion criteria, the clinical research team prepared the TD-NIRS system in the neuro-interventional radiology unit. Once the patient was transferred to the operating table and intubated, TD-NIRS probes were attached to the patient’s forehead using a medical-grade adhesive (Figure 2A). To avoid any delays in door-to-puncture time, probe placement was performed immediately after intubation and while the patient was being prepped for EVT. The probes were strategically positioned to measure the MCA superior division vascular territory on both hemispheres, with a 3 cm separation between each optical source and detector (Figure 2B-D). The placement of the radio-opaque TD-NIRS probes and optical fibers was arranged to minimize interference with fluoroscopy imaging throughout the procedure (Figure 2). TD-NIRS measurements were taken every 20 seconds over the monitoring period, with all patients measured for a minimum of 7 minutes and 20 seconds centered at the recanalization time. These TD-NIRS measurements began at the start of the EVT procedure and continued until after recanalization. To account for local changes during clot removal—including blood flow reversal, inflation of balloon guiding catheter, vessel spasm, and vessel collapse—the recanalization window, defined as the two-minute period centered at recanalization time, was excluded from analysis. The pre- and post-recanalization measurement windows, which were used in all analyses, were defined as the 2 minute and 20-40 second period immediately before and after recanalization window. This yielded 17-18 total TD-NIRS measurements, with 9 from the pre-recanalization period and 8-9 from the post-recanalization period. Each TD-NIRS measurement provided a 10.24 ns time-domain trace, resolved into 1024 data points corresponding to 10 ps resolution. The initial and final 200 time points were excluded to isolate the relevant signal, which begins to rise after time point 200 and fully decays before time point 800, resulting in a 6 ns duration TD-NIRS trace for analysis. Throughout the EVT procedure, vital parameters, including BP, HR, SpO_2_, fraction of inspired oxygen (FiO_2_), and end-tidal carbon dioxide (ETCO_2_), were continuously recorded.

**Figure 2.**
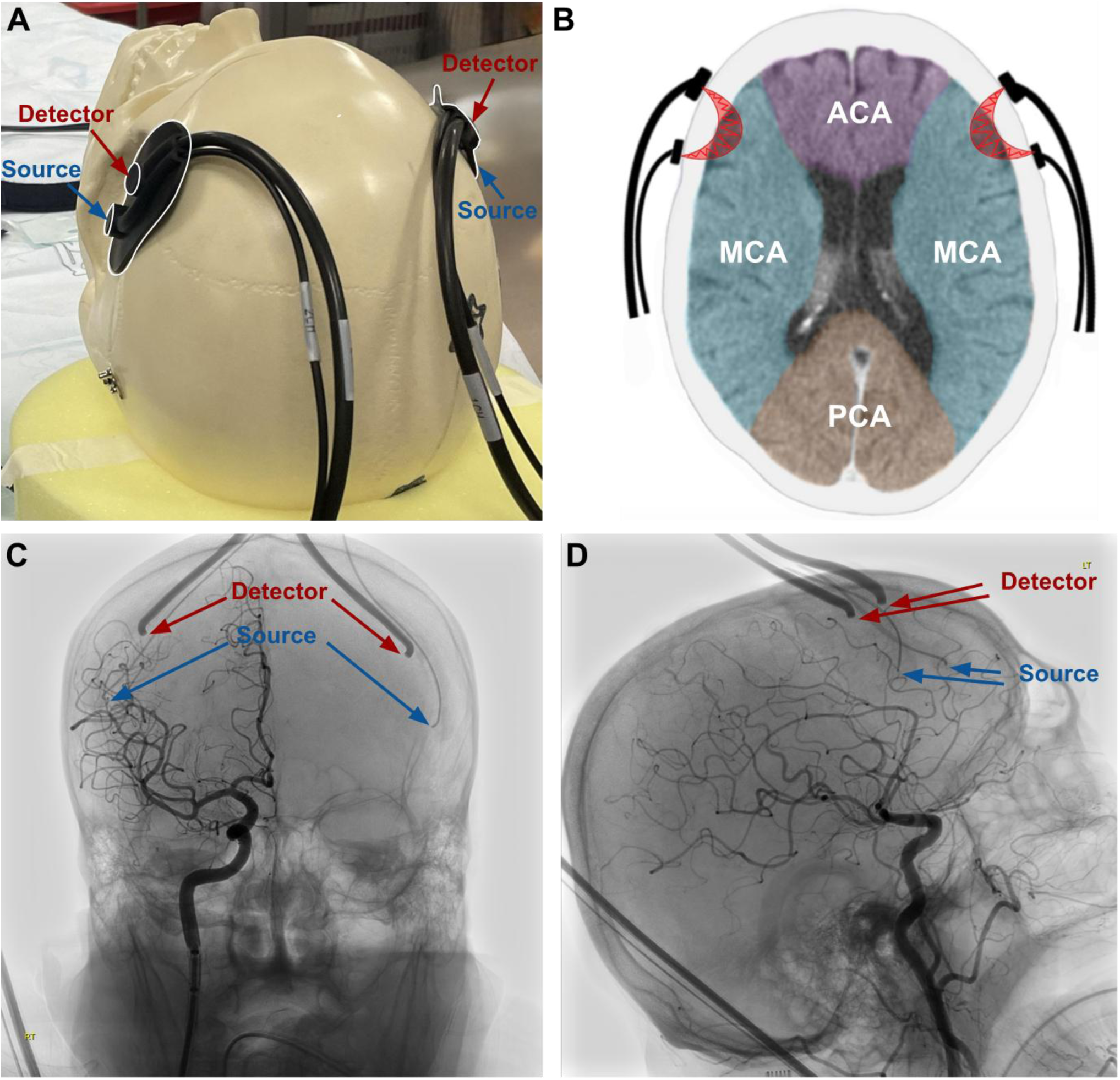
Placement of TD-NIRS Probes on the Scalp for Brain Monitoring. (A) TD-NIRS probes are placed on the forehead using a medical-grade adhesive to measure the MCA superior division territories. (B) Axial head CT scan illustrating TD-NIRS source and detector probe placement relative to MCA superior division territories; ACA, MCA, and PCA territories are shaded in purple, blue, and orange, respectively. (C) Anterior-posterior fluoroscopy image displaying TD-NIRS probe positioning, capturing bilateral MCA superior division territories, with right MCA territory visualized by contrast injection via the right internal carotid artery. (D) Lateral fluoroscopy image showing TD-NIRS probes on both hemispheres, with right MCA territory visualized by contrast injection in the right internal carotid artery. TD-NIRS, Time-Domain Near-Infrared Spectroscopy; CT, Computed Tomography; MCA, Middle Cerebral Artery; ACA, Anterior Cerebral Artery; PCA, Posterior Cerebral Artery.

After recording, the raw TD-NIRS data was checked for noise artifacts, low signal-to-noise ratios, and signal shifts. Following data validation, the TD-NIRS data was analyzed using a transport-rigorous sMC approach with a reference simulation generated by our open-source conventional Monte Carlo (cMC) simulation engine to determine [HbO_2_] and [HHb].(31,34) Specifically, the sMC approach rapidly computes predictions of TD-NIRS signals from a specified set of scattering coefficients and concentrations of optical absorbers. The sMC analysis was utilized in conjunction with a non-linear optimization algorithm to simultaneously determine the wavelength-dependent reduced scattering coefficient *μ*′_s_ using a “power-law” spectral constraint for scattering as well as [HbO_2_] and [HHb].(35,36) To our knowledge, while proposed by other investigators, this transport-rigorous sMC method has not been implemented to analyze TD-NIRS signals in the brain and represents a more accurate alternative to the standard diffuse approximation commonly used for analyzing TD-NIRS signals.(25) Thus, the sMC approach provides a more robust and accurate prediction of TD-NIRS signals and recovery of physiological properties.(25,31,36,37) While cMC methods incur high computational cost, the sMC method exploits scaling relations from a single pre-executed cMC simulation.(31,38) In this study, we examine the use of an sMC inverse model to provide accurate predictions of cerebral optical and hemodynamic properties.

### Statistical Analysis

MATLAB, R Studio, SPSS, and GraphPad software were employed to combine, analyze, and plot the optical parameters of the sMC-based TD-NIRS data—particularly [HbO_2_], [HHb], [tHb], and StO_2_—alongside clinical data—including BP, HR, ETCO_2_ and SpO_2_. The Wilcoxon matched pairs signed-rank exact test was employed to compare sMC processed TD-NIRS parameters pre- and post-recanalization, and the median (x̃) and interquartile range (IQR) of the paired differences was reported. Mann-Whitney U test was utilized to compare TD-NIRS derived Δ[HbO_2_]/[HHb] in patients who developed post-EVT cortical infarct with those who did not, and median and IQR were reported for each group. Due to the small sample size, the data set was not assumed to be normally distributed, and non-parametric tests were selected. The effect size (*r*) was also reported. Additionally, Firth’s logistic regression and ROC analysis were conducted to evaluate Δ[HbO_2_]/[HHb] as a predictor of post-EVT infarct. For these analyses, patients were dichotomized into yes/no infarct conditions, as detected on post-EVT MRI in the territory detected by TD-NIRS. Standard error was the method of variance for the ROC analysis. Regression inputs were scaled, and the odds ratio (OR) was reported for Firth’s logistic regression analysis.(39) Statistical significance was defined as *p*<0.05. Moving averages using 1-minute windows were applied to all linear representations of sMC processed TD-NIRS data, which were utilized for visualization purposes. Sex was not included as a covariate in analysis because Mann-Whitney U test revealed no significant differences between males and females pre- vs post-recanalization in [HbO_2_], [HHb], [tHb], StO_2_ on the affected side (*p*=0.41, 0.32, 0.65, 0.53, respectively), unaffected side (*p*=0.41, 1.0, 0.23, 0.32, respectively), the affected/unaffected ratio (*p*=0.55, 0.53, 0.41, 0.23, respectively), or the Δ[HbO_2_]/[HHb] (*p*=0.53) (*n*=11).

## Results

### Demographics and Clinical Characteristics of Patients

The study included eleven patients enrolled over an 18-month period who underwent EVT and TD-NIRS monitoring. Demographics and clinical characteristics of the cohort are presented in Tables 1 and 2, respectively. The average times of major events preceding thrombectomy are listed in Table S1. The median (IQR) arterial blood hemoglobin level was 13 (12–14) g/dL pre-EVT and 12 (11–14) g/dL post-EVT, indicating no considerable blood loss or hemodilution during EVT. Respiratory and hemodynamic conditions (MAP, SpO_2_, ETCO_2_, and FiO_2_) were stable throughout the procedure (Table S2).

**Table 1.** Demographics of TD-NIRS-Recorded Patients (*n*=11) Undergoing Thrombectomy.

|  |  |
| --- | --- |
| Age -median (IQR) | 73 (70-84) |
| Male Sex - number (%) | 4 (36%) |
| Female Sex - number (%) | 7 (64%) |
| Race or Ethnic Group - number (%) |  |
| American Indian or Alaska Native | 0 (0%) |
| Asian | 7 (64%) |
| Black or African American | 1 (9%) |
| Hispanic or Latino | 0 (0%) |
| White | 2 (18%) |
| Other Race or Mixed Race | 1 (9%) |
| Risk Factors - number (%) |  |
| Hypertension | 9 (82%) |
| Hyperlipidemia | 7 (64%) |
| Diabetes | 4 (36%) |
| Heart Disease | 6 (55%) |
| Smoking |  |
| Yes | 0 (0%) |
| No | 6 (55%) |
| Unknown | 5 (45%) |
| Former | 0 (0%) |
| Atrial Fibrillation | 8 (73%) |
| Prior stroke or TIA | 3 (27%) |
| Intracranial Atherosclerosis | 8 (73%) |

Abbreviations: IQR, interquartile range; TIA, transient ischemic attack.

**Table 2.**
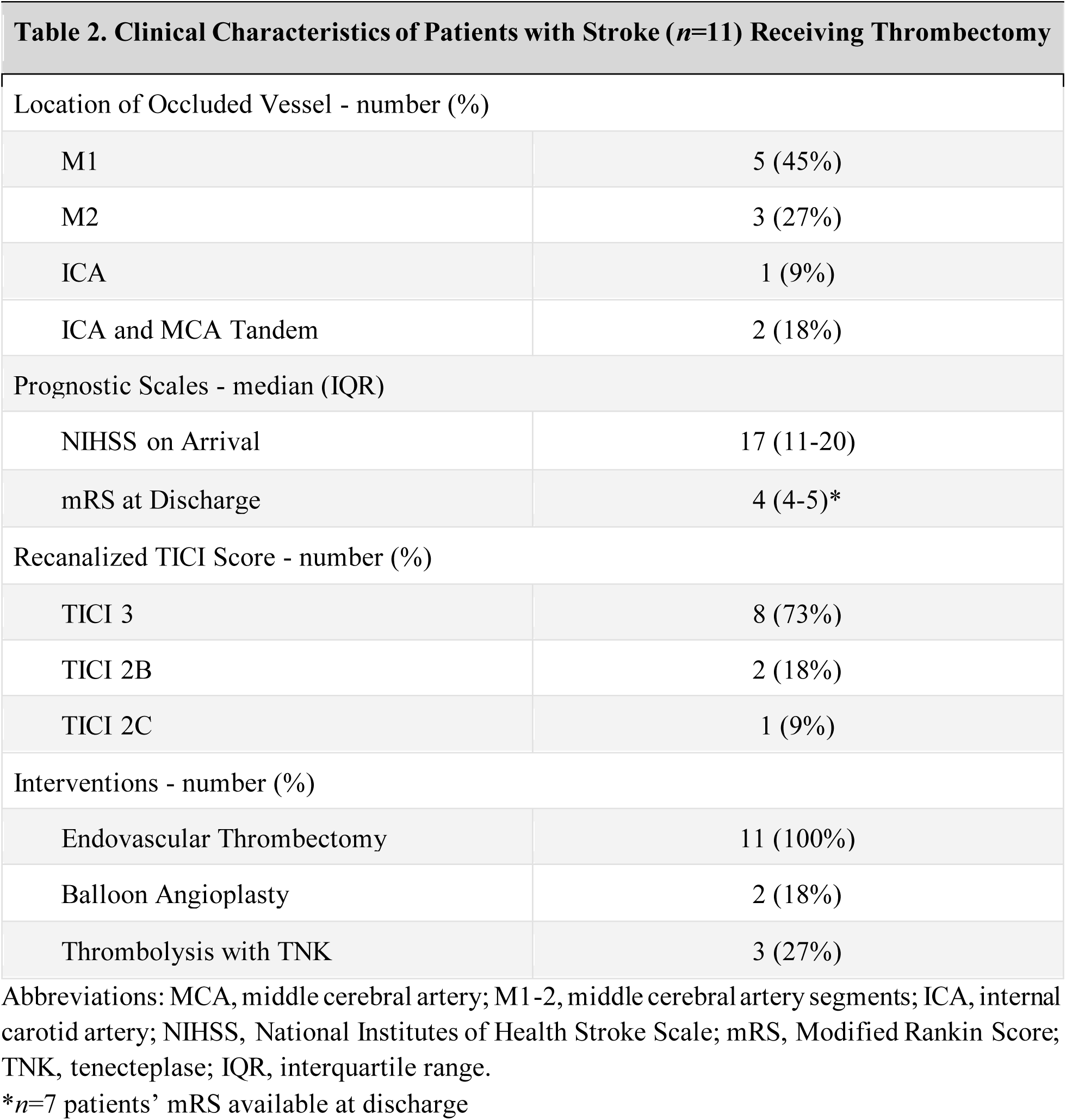
Clinical Characteristics of Patients with Stroke (*n*=11) Receiving Thrombectomy.

|  |  |
| --- | --- |
| Location of Occluded Vessel - number (%) |  |
| M1 | 5 (45%) |
| M2 | 3 (27%) |
| ICA | 1 (9%) |
| ICA and MCA Tandem | 2 (18%) |
| Prognostic Scales - median (IQR) |  |
| NIHSS on Arrival | 17 (11-20) |
| mRS at Discharge | 4 (4-5)* |
| Recanalized TICI Score - number (%) |  |
| TICI 3 | 8 (73%) |
| TICI 2B | 2 (18%) |
| TICI 2C | 1 (9%) |
| Interventions - number (%) |  |
| Endovascular Thrombectomy | 11 (100%) |
| Balloon Angioplasty | 2 (18%) |
| Thrombolysis with TNK | 3 (27%) |

Abbreviations: MCA, middle cerebral artery; M1 -2, middle cerebral artery segments; ICA, internal carotid artery; NIHSS, National Institutes of Health Stroke Scale; mRS, Modified Rankin Score; TNK, tenecteplase; IQR, interquartile range.
\* $n=7$ patients' mRS available at discharge

### Pre- and Post-Endovascular Thrombectomy Imaging Results

Pre-EVT head CT scans were assessed for vascular occlusions and the extent of cerebral infarct. All patients presented with consequential occlusions in the MCA or ICA territory, involving the superior MCA division. Successful reperfusion was achieved in all patients, with eight patients attaining TICI 3 score, two patients attaining TICI 2b score, and one patient attaining a TICI 2c score (Table S3). Post-EVT MRI scans were obtained to evaluate the effectiveness of reperfusion and identify new infarcts. Despite achieving TICI scores of 2b-3, six patients’ post-EVT MRI demonstrated cortical infarct in TD-NIRS measurement territories (Table S3).

### Real-Time TD-NIRS Monitoring during Endovascular Thrombectomy

Figures 3A and 3B present perioperative neuromonitoring timelines alongside neuroimaging data, including pre-EVT head CT, intra-EVT fluoroscopy images, and post-EVT MRI for two representative patients from the cohort (patient 1, Figure 3A; and patient 7, Figure 3B). Real-time TD-NIRS measurements from the affected side, along with MAP (via arterial line), are illustrated as linear graphs. As depicted in Figure 3A (patient 1), [HbO_2_], [tHb], and StO_2_ all increased immediately after the vascular recanalization window, which was confirmed with a TICI score of 3. This underscores the robustness of TD-NIRS for real-time monitoring of brain tissue oxygenation during reperfusion. Figure 3B shows another case (patient 7) achieving a TICI 2b score after successful recanalization of the MCA inferior division. However, despite several attempts, the MCA superior division—corresponding to vascular territory beneath the TD-NIRS probe—remained occluded. Notably, the TD-NIRS optical parameters remained stable throughout the procedure, likely due to failed recanalization. The linear graphs of TD-NIRS measurements along with continuous MAP depicting unaffected sides for both patients are presented in Figure S1.

**Figure 3.**
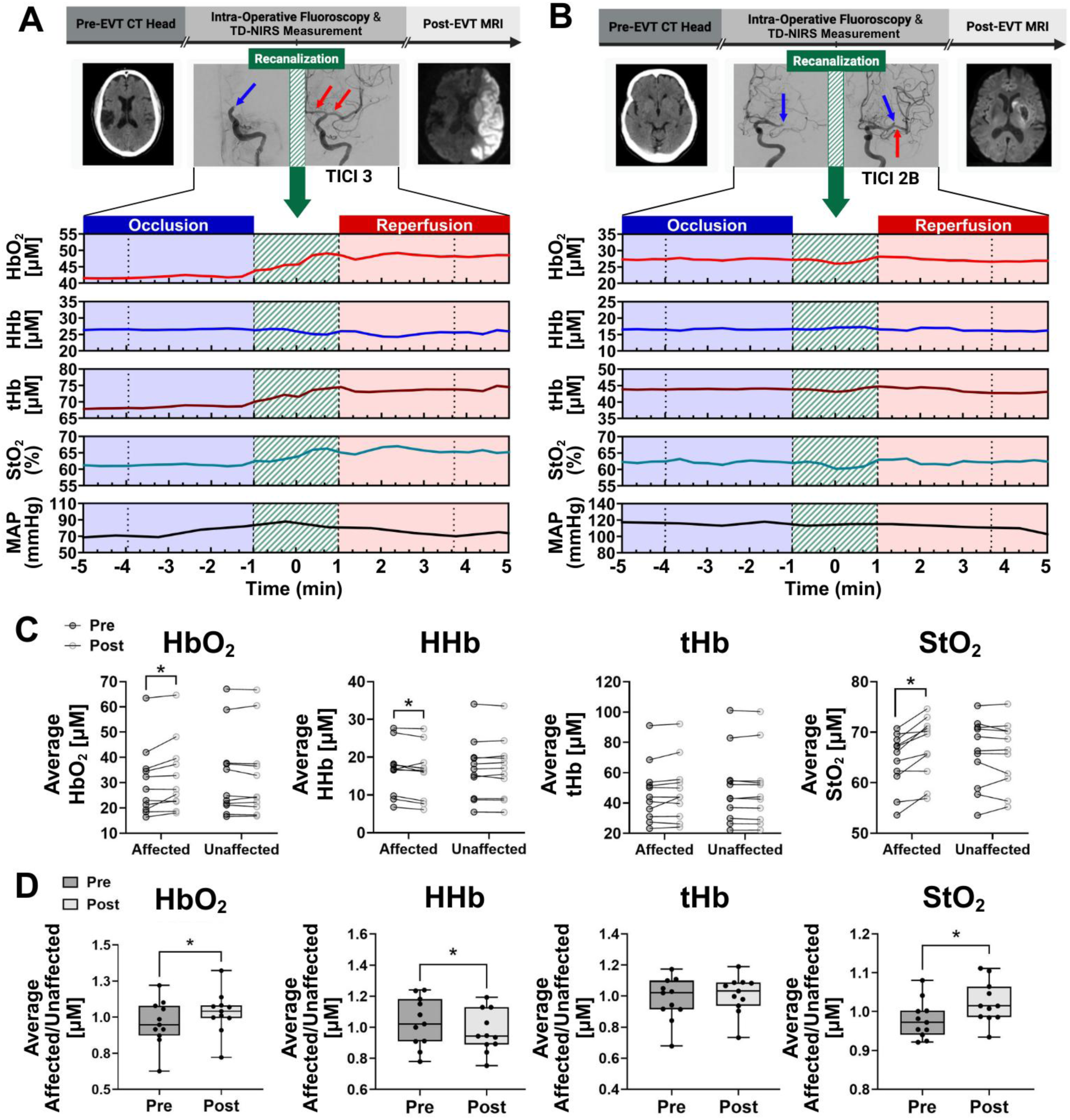
Perioperative neuroimaging and optical neuromonitoring of two representative patients during endovascular thrombectomy (EVT), as well as cumulative pre- and post-EVT TD-NIRS values across eleven patients. (A and B) Neuromonitoring sequences from two representative cases (A: Patient 1; B: Patient 7), including pre-EVT non-contrast head CT, intra-EVT fluoroscopy (blue arrows denote occluded vessels; red arrows denote recanalized vessels), and post-EVT MRI. Time series of TD-NIRS and blood pressure values during EVT display [HbO_2_], [HHb], [tHb] concentrations (µM), StO_2_ (%), and MAP (mmHg). Dotted lines indicate the 2.5-minute pre- and post-recanalization data intervals used for analysis. (C) Wilcoxon matched-pairs signed-rank exact test reveals significant changes between the 2.5 minutes pre and post on the affected side for [HbO_2_] (x̃=1.5 [IQR: 0.19 - 4.1], *r*=0.78, *p*=0.0068), [HHb] (x̃=−0.86 [IQR: −1.3 - −0.16], *r*=−0.62, *p*=0.042), and StO_2_ (x̃=3.4 [IQR: 0.71 - 4.3], *r*=0.86, *p*=0.0020), while [tHb] showed no significant change (x̃=0.97 [IQR: −0.39 - 2.1], *r*=0.46, *p*=0.15). No significant changes were observed on the unaffected side ([HbO_2_]: x̃=−0.22 [IQR: −0.83 - 0.54], *r*=−0.21, *p=*0.52; [HHb]: x̃=−0.049 [IQR: −0.23 - 0.33], *r*=0.11, *p=*0.76; [tHb]: x̃=−0.27 [IQR: −0.76 - 0.11], *r*=−0.24, *p=*0.46; StO_2_: x̃=−0.084 [IQR: −1.3 - 0.35], *r*=−0.13, *p=*0.70) (*n*=11). (D) Box plots illustrate the affected/unaffected ratio of 2.5 minute average TD-NIRS-derived values pre- and post-recanalization, with Wilcoxon matched-pairs signed-rank exact tests confirming significant differences in [HbO_2_] (x̃=0.081 [IQR: 0.037 - 0.10], *r*=0.75, *p*=0.0098), [HHb] (x̃=−0.053 [IQR: −0.13 - −0.016], *r*=−0.67, *p*=0.024), and StO_2_ (x̃=0.047 [IQR: 0.024 - 0.062], *r*=0.83, *p*=0.0029). No significant differences were observed in [tHb] (x̃=0.023 [IQR: −0.013 - 0.039], *r*=0.59, *p*=0.054) (*n*=11). Bars represent the range of dataset. TD-NIRS, time-domain near-infrared spectroscopy; CT, computed tomography; MRI, magnetic resonance imaging; [HbO_2_], oxyhemoglobin concentration (µM); [HHb], deoxyhemoglobin concentration (µM); [tHb], total hemoglobin concentration (µM); StO_2_, tissue oxygen saturation (%); MAP, mean arterial pressure (mmHg); * indicates *p* < 0.05; (µM) = micromolar.

To assess the immediate changes in cerebral oxygenation following recanalization, the Wilcoxon matched pairs signed-rank exact test was employed. This test compared the mean 2.5-minute TD-NIRS measurements pre- and post-recanalization window on the affected and unaffected sides across all eleven patients (Figure 3C). On the affected side, notable differences were observed between pre- and post-recanalization values for [HbO_2_] (x̃=1.5 [IQR: 0.19 - 4.1], *r*=0.78, *p*=0.0068), [HHb] (x̃=−0.86 [IQR: −1.3 - −0.16], *r*=−0.62, *p*=0.042), and StO_2_ (x̃=3.4 [IQR: 0.71 - 4.3], *r*=0.86, *p*=0.0020). In contrast, these parameters on the unaffected side did not show prominent differences, with *p*-values of 0.52, 0.76, and 0.70, respectively. No noteworthy differences were observed for [tHb] values on both the affected (*p*=0.15) and unaffected (*p*=0.46) sides.

Additionally, Wilcoxon matched pairs signed-rank exact test was utilized to examine relative oxygenation changes by comparing affected-to-unaffected side ratio (affected/unaffected) of the mean 2.5-minute TD-NIRS measurements pre- and post-recanalization across all eleven patients (Figure 3D). Significant differences were observed between pre- and post-recanalization affected/unaffected values for [HbO_2_] (x̃=0.081 [IQR: 0.037 - 0.10], *r*=0.75, *p*=0.0098), [HHb] (x̃=−0.053 [IQR: −0.13 - −0.016], *r*=−0.67, *p*=0.024), and StO_2_ (x̃=0.047 [IQR: 0.024 - 0.062], *r*=0.83, *p*=0.0029). The [tHb] value pre- and post-recanalization showed no notable difference (x̃=0.023 [IQR: −0.013 - 0.039], *r*=0.59, *p*=0.054).

### Comparison of Pre- and Post-Recanalization HbO_2_/HHb Ratio and Its Association with Post-EVT Brain Infarct

When comparing the difference between pre- and post-recanalization [HbO_2_]/[HHb] ratios (Δ[HbO_2_]/[HHb]) on the affected side, we observed a significant increase in Δ[HbO_2_]/[HHb] in patients who subsequently developed cortical infarction on post-EVT MRI. In contrast, patients who did not develop post-EVT cortical infarct showed little or no change in Δ[HbO_2_]/[HHb] (Figure 4C). To further illustrate this relationship, we present linear graphs of changes in [HbO_2_] and [HHb] on both affected and unaffected sides from two representative cases. In the first case (Figure 4A; patient 1), who developed a post-thrombectomy cortical infarct, relative hemoglobin (rHb) on the affected side increased markedly compared to pre-recanalization. In contrast, the second case (Figure 4B; patient 11), who did not develop a cortical infarct, showed relatively stable rHb levels.

**Figure 4.**
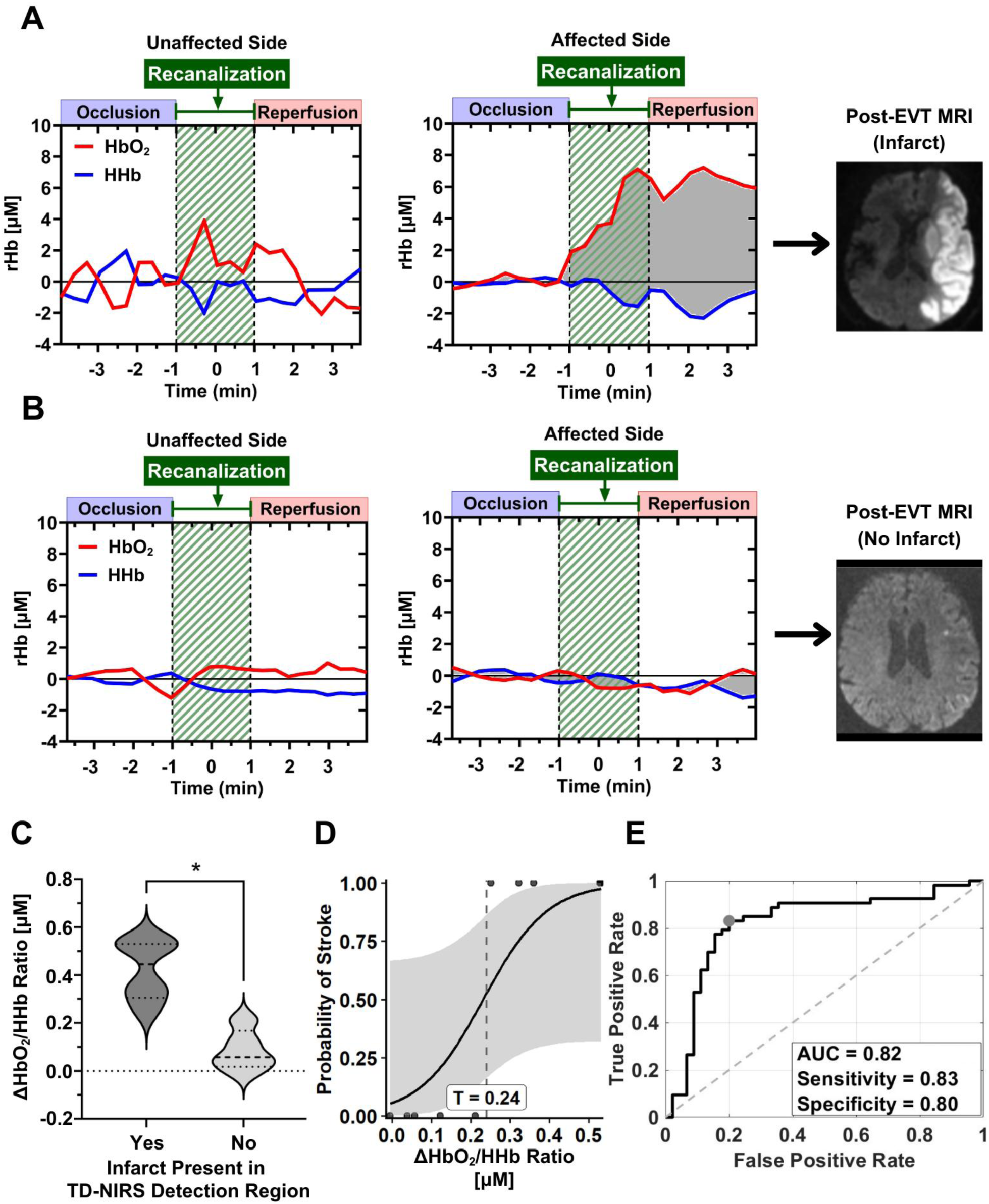
Pre- and Post-EVT ΔHbO_2_/HHb Ratio Variations Linked to Post-EVT Cortical Infarct. (A and B) Line graphs of relative TD-NIRS optical parameters ([HbO_2_] and [HHb]) pre-and post-recanalization on both unaffected and affected sides, as well as post −EVT diffusion-weighted MRI images, for two representative cases, patient 1 and patient 11, that achieved TICI 3 reperfusion. (C) Distribution of the average 2.5-minute Δ[HbO_2_]/[HHb] in patients with and without post-EVT cortical infarction in the TD-NIRS detection region. Mann-Whitney U test revealed notable differences in the aforementioned parameter (x̃=0.44 [IQR: 0.30 - 0.53] vs x̃=0.058 [IQR: 0.017 - 0.17], *r*=−0.83, *p*=0.0043, *n*=11). (D) Firth’s logistic regression shows the predictive strength of 2.5-minute Δ[HbO_2_]/[HHb] for post-EVT cortical infarction (OR=3.8, *p*=0.0041, *n*=11). *T*=0.24 indicates the threshold for infarct prediction, determined through ROC analysis. (E) ROC curve analysis demonstrates the predictive capability of Δ[HbO_2_]/[HHb] for post-EVT cortical infarct, with 0.24 being the optimal threshold for prediction (AUC: 0.82, 95% CI: [0.71 - 0.90], Sensitivity: 0.83, Specificity: 0.80, *n*=11). [rHb], relative hemoglobin concentration (µM); [HbO_2_], oxyhemoglobin concentration (µM); [HHb], deoxyhemoglobin concentration (µM); EVT, endovascular thrombectomy; MRI, magnetic resonance imaging; TD-NIRS, time-domain near-infrared spectroscopy; *indicates *p* < 0.05; (µM) = micromolar

To assess statistical significance, Mann-Whitney U test was conducted on all eleven patients in the cohort to assess the Δ[HbO_2_]/[HHb] ratio pre- and post-recanalization (Figure 4C). This comparison was made between patients with and without cortical infarct in the respective vascular territory, as detected on post-EVT MRI. We defined the respective vascular territory as the cortical region directly beneath the placement of the TD-NIRS probe, encompassing the area detectable by the TD-NIRS sensors on either side (Figure 2B). The results showed a notable difference in Δ[HbO_2_]/[HHb] ratio between patients with and without cortical infarct (x̃=0.44 [IQR: 0.30 - 0.53] vs x̃=0.058 [IQR: 0.017 - 0.17], *r*=−0.83, *p*=0.0043, *n*=11) (Figure 4C). This result demonstrates that a higher [HbO_2_]/[HHb] ratio post-recanalization, compared to pre-recanalization, is strongly associated with cortical infarction post-EVT.

We employed Firth’s logistic regression to further evaluate the predictive value of the TD-NIRS-derived Δ[HbO_2_]/[HHb] ratio for detecting post-EVT cortical infarction (Figure 4D). The model, which compared the [HbO_2_]/[HHb] ratio difference before and after recanalization, yielded a logistic regression classifier that performed better than chance (OR=3.8, *p*=0.0041, *n*=11) (Figure 4D). A threshold value of *T*=0.24 for infarct prediction was determined through ROC analysis. These results suggest that the changes in the [HbO_2_]/[HHb] ratio immediately after recanalization may serve as a meaningful marker to predict the likelihood of developing post −EVT cortical infarction.

Finally, we assessed the ability of Δ[HbO_2_]/[HHb] to predict post-EVT cortical infarction using receiver operating characteristic (ROC) curve analysis (Figure 4E). The ROC model evaluates the ability of Δ[HbO_2_]/[HHb] ratio to classify infarct vs. non-infarct outcomes. The results indicate strong predictive performance, with an optimal threshold of 0.24 (AUC=0.82, 95% CI [0.71 - 0.90]), yielding a sensitivity of 0.83 and specificity of 0.80. The high AUC suggests that Δ[HbO_2_]/[HHb] has strong potential to serve as a reliable biomarker for identifying patients at risk of cortical infarction post-EVT.

## Discussion

Our results demonstrate the ability of TD-NIRS to provide continuous, real-time, non-invasive measurements of cerebral optical properties throughout the EVT procedure and the potential to identify patients at risk of post-EVT cortical infarct. As shown in Figure 3A, during the recanalization period in patient 1, intraoperative TD-NIRS measurements capture a marked change in tissue optical properties—[HbO_2_], [HHb], and StO_2_—immediately following successful reperfusion (TICI 3). This highlights the sensitivity of the TD-NIRS system in capturing real-time, tissue-level dynamic changes that reflect restored cerebral perfusion in response to EVT. These results are supported by significant increases seen in [HbO_2_], [HHb], and StO_2_ on the affected side post-recanalization (Figure 3C) and notable changes in affected/unaffected ratio for the same parameters post-recanalization (Figure 3D), underscoring the potential of TD-NIRS to detect and monitor cerebral tissue oxygenation changes during EVT. Comparing the affected and unaffected hemispheres provides an internal baseline control that helps account for potential systemic physiological fluctuations—such as changes in blood pressure, variations in FiO_2_—that may influence cerebral blood flow and oxygenation bilaterally. By minimizing changes relative to the unaffected side, we aim to isolate localized perfusion changes attributable to successful recanalization. This method allows for a more focused assessment of the sensitivity and specificity of the TD-NIRS system in detecting rapid, cerebral perfusion-related changes in the affected hemisphere immediately following EVT.

In contrast, individuals such as patient 7, who did not achieve successful recanalization in the MCA superior division—the vascular territory directly beneath the TD-NIRS probes—had optical parameters that remained stable (Figure 3B). Although this patient attained partial recanalization in the MCA (TICI 2b), this area was outside the TD-NIRS detection region. The lack of notable change in TD-NIRS parameters following partial recanalization demonstrates TD-NIRS’ specificity in detecting tissue oxygenation changes within its monitoring region (Figure 3B). It’s important to highlight that despite the failed recanalization, patient 7 did not develop a subsequent infarction in the territory supplied by the occluded MCA superior division. This outcome may reflect effective collateral circulation, allowing maintenance of tissue viability despite the absence of reperfusion. Notably, prior studies have shown that patients with similar baseline characteristics and TICI scores can still have different outcomes, reflected by variation in the mRS. This variability is often attributed to individual differences in collateral circulation, a critical predictor of stroke outcomes, irrespective of a fixed time window.(40,41) Therefore, an individualized approach that employs a multimodal monitoring system capable of assessing perfusion and metabolism may help optimize patient outcomes. Future studies should examine whether TD-NIRS-derived optical signatures correlate with validated quantitative measures of collateral status, such as the ASITN/SIR collateral grading scale or automated collateral scoring on CTA.(42,43) Although TICI 2b-3 scores are generally considered indicators of successful reperfusion, growing evidence suggests that not all TICI scores confer equal clinical benefit. Generally, patients achieving TICI 3 reperfusion tend to have significantly better outcomes compared to those with TICI 2b or 2c.(44) While our current results from 11 patients did not show a significant difference between TICI 2b and TICI 3, they support the feasibility and potential of TD-NIRS to fill this gap by providing real-time cerebral tissue oxygenation. This highlights a critical gap in current intra-procedural assessment tools, which lack the ability to determine, in real-time, whether the achieved reperfusion is sufficient for a given patient. A future large-scale study with more granular patient stratification is warranted to evaluate whether TD-NIRS can help identify which patients benefit from TICI 2b and which require further thrombectomy attempts to achieve TICI 3, enabling more personalized and outcome-driven treatment decisions.

Furthermore, despite achieving optimal reperfusion as indicated by TICI scores of 2b and 3, our study reveals variability in neurological outcomes, as quantified by post-thrombectomy brain MRI. Notably, 6 out of 11 patients suffered from post-thrombectomy cortical infarct that were detected on MRI scans, despite achieving high TICI scores (Table S3). These post −EVT MRI results highlight the disconnect between TICI scores and clinical outcomes, suggesting that the TICI grading system may not be a reliable indicator of post-reperfusion brain tissue viability. This observation is supported by a meta-analysis of clinical studies, which reported substantial infarct growth despite successful reperfusion, defined by TICI score of 2b-3.(45) A recent review on beneficial versus futile and harmful reperfusion emphasizes that angiographic recanalization does not guarantee tissue recovery, highlighting the importance of early identification of post −EVT cortical infarct.(46)

A particularly compelling finding in our study is the difference in Δ[HbO_2_]/[HHb]—the ratio of oxygenated (supplied) to deoxygenated (metabolized) hemoglobin—before and after recanalization. As shown in Figure 4C, patients who developed post-EVT cortical infarct within 24-48 hours demonstrate a significantly greater increase in this ratio compared to those without infarction. These findings suggest that the magnitude of change in cerebral oxygen supply ([HbO_2_]) and consumption ([HHb]), a marker of metabolism by live tissue, relative to pre-EVT, may serve as an early biomarker of tissue injury despite angiographic success. This observation supports the hypothesis that a mismatch between cerebral tissue perfusion and metabolism during the reperfusion period is strongly associated with the extent of brain tissue infarction, particularly in patients with severe brain injury, potentially due to absent or inadequate leptomeningeal collateral circulation. These findings are corroborated by Firth’s logistic regression (Figure 4D) and ROC (Figure 4E) analyses that show Δ[HbO_2_]/[HHb] to be significantly predictive of post-EVT cortical infarct. This pattern is further illustrated by two representative cases shown in Figures 4A and 4B. Despite achieving TICI 3 reperfusion status, both patients (patient 1 and patient 11) had drastically different clinical outcomes. In patient 1, who developed cortical infarct on follow-up MRI, the linear TD-NIRS graph demonstrates a marked difference between [HbO_2_] and [HHb] immediately post-recanalization relative to the pre-recanalization period (Figure 4A). In contrast, patient 11, who did not develop cortical infarct, shows relatively stable trends, with a smaller gap between relative [HbO_2_] and [HHb] (Figure 4B). These findings indicate that the TICI score alone may not provide comprehensive predictive value regarding the infarct progression and overall patient outcomes—underscoring the need for a more nuanced assessment of cerebral oxygenation, metabolism, and tissue viability post-EVT.(19,47,48) The introduction of Δ[HbO_2_]/[HHb] as a physiological biomarker has potential clinical value and may complement angiographic assessments by revealing the extent of reperfusion at the tissue level—providing critical information to help interventionalists determine whether further thrombectomy attempts may be beneficial. While the underlying mechanisms remain to be fully elucidated, these findings highlight the potential of TD-NIRS-derived Δ[HbO_2_]/[HHb] to serve as a surrogate marker for tissue viability during EVT, with implications beyond the procedural setting into neurocritical care management. Post-EVT care, such as systemic blood pressure management, is a highly debated and individualized challenge.(49,50) In current practice, decisions regarding BP targets are often made without direct insight into the brain’s perfusion and metabolism status and whether tissue remains viable post-thrombectomy. Early biomarkers like Δ[HbO_2_]/[HHb] could support patient-specific bedside BP management—allowing clinicians to avoid cerebral hypoperfusion from overly aggressive BP reduction or prevent reperfusion injury and hemorrhagic transformation from permissive hypertension. Ultimately, these findings suggest that monitoring changes in Δ[HbO_2_]/[HHb] could be crucial in identifying patients at risk for expansion of infarct or reperfusion injury following the procedure.

Additionally, most current TD-NIRS systems utilize a homogeneous tissue model for data analysis, which may result in less accurate cerebral tissue oxygenation metrics due to the composite nature of optical signals from both the cerebral and extracerebral layers. This limitation underscores the need for advanced multilayer optical analysis and modeling to better distinguish and interpret signals from discrete tissue layers, particularly in cerebral perfusion and hemodynamic assessments. Future directions should focus on refining the TD-NIRS system to improve its ability to accurately measure cerebral oxygenation. These advancements could include the integration of more sophisticated models and algorithms capable of differentiating between extracerebral and cerebral signals, thereby improving the precision of the system in clinical settings.

Finally, future studies in larger cohorts will be essential to account for patient-specific variability, including comorbidities, collateral status, infarct size, and infarct evolution. Achieving this level of evidence remains challenging due to logistical constraints, particularly the need for timely informed consent prior to thrombectomy in an emergency setting. Broader adoption of Exception from Informed Consent protocols or FDA approval of the device could help facilitate more rapid enrollment and accelerate high-quality data acquisition necessary for clinical integration. In addition, exploring the role of bedside TD-NIRS monitoring in the ICU (post-EVT) is warranted to assess its utility in detecting delayed hypoperfusion, reperfusion injury, or secondary infarct progression.

### Study Limitations

This study has several limitations. The sample size was relatively small, which limits statistical power and generalizability of the findings, emphasizing the need for larger multi-center studies. The current TD-NIRS system was limited to monitoring only the MCA territory to avoid interference with anteroposterior and lateral fluoroscopic visualization during the EVT procedure. As a result, other vascular territories, including inferior division of MCA, as well as anterior and posterior cerebral artery territories, could not be monitored. Despite these limitations, it is important to note that no adverse effects related to TD-NIRS monitoring were observed, and angiographic visualization was not compromised, as the radio-opaque probes were deliberately positioned to avoid overlap with critical neuroimaging planes. Additionally, potential confounders such as collateral status and infarct volume were not controlled or adjusted for. Future studies with broader probe configurations and better patient stratification are warranted to refine its clinical applicability.

## Conclusion

This study demonstrates the potential of TD-NIRS as a real-time, non-invasive tool for continuous monitoring of cerebral microvascular oxygenation and hemodynamics during EVT. Additionally, the relationship between post-EVT outcomes and optical parameters, independent of reperfusion (TICI) scores, suggests that TD-NIRS could play a critical role in assessing brain tissue viability immediately after recanalization and guide intraoperative decision-making and post-EVT management. The observed trends in pre- versus post-recanalization [HbO_2_]/[HHb] ratios highlight the potential clinical relevance of these measurements in predicting post −EVT brain infarction. While further research involving larger cohorts and advanced analytical techniques is needed, integrating TD-NIRS measurement with transport-rigorous analysis for accurate spectral recovery of tissue scattering and blood concentration parameters into EVT protocols may improve the precision of neuro-interventional procedures, leading to better patient outcomes and more comprehensive monitoring during EVT. Future directions should focus on refining TD-NIRS measurement and data analysis techniques, expanding patient cohorts, and developing a multimodal monitoring approach integrating both cerebral perfusion and metabolism during and after EVT.

## Acknowledgements

The authors are grateful to the neurocritical care research team at the Akbari Lab—Samantha Le Phan, Marissa St. Marseille, Michael Robles, Jessica Le, Yeayoung Vac, Doyi Moon, Kayla Langis, Siddharth Karthikeya, Sarina Rai, Anushka Singhal, and Tatiana To—for their contributions to patient screening and data collection. The authors also thank Dr. Bernard Choi (University of California, Irvine, Departments of Surgery and Biomedical Engineering) for administrative support and resource allocation and Dr. Michael Helton (University of California, Irvine, Beckman Laser Institute & Medical Clinic) for contributions to sMC analysis and modeling. Appreciation is extended to the neurointerventional radiology and neuro-ICU teams, including nurses and technicians, at the UC Irvine Medical Center, for their support with patient enrollment and their cooperation and flexibility throughout the study.

## Statements and Declarations

### Ethical Considerations

This study was approved by the Institutional Review Board at the University of California, Irvine (IRB#20195366).

### Consent to Participate

Written informed consent was obtained from the legally authorized representatives of all patients.

### Consent for Publication

Written informed consent was obtained from the legally authorized representatives of all patients, including consent for publication.

### Data Availability Statement

The datasets generated during and/or analyzed during the current study are available in the Mendeley Data repository, DOI: 10.17632/2kk6p4kgyk.1

## Supplemental Material

The following supplemental material can be found on the online version of this manuscript:

Tables S1-S3

Figure S1

Figure S2

