## Supplemental Material for "Continuous Time-Domain Near-Infrared Spectroscopy During Endovascular Thrombectomy Enables Early Prediction of Post-Recanalization Cortical Infarct"

| Table S1. Average Times of Major Events Prior to Endovascular Thrombectomy (h:mm) | |
|---|---|
| Average time from LKW to CT w/o contrast time | 6:17 |
| Average time from LKW to TNK injection | 1:56* |
| Average time from TNK injection to groin puncture time | 0:46* |
| Average time from CT w/o contrast to puncture time | 0:35 |
| Average duration of TD-NIRS recording | 1:12 |
| Average stroke notification to consent time | 0:38 |
| Average stroke notification to TD-NIRS recording time | 0:49 |

**n*=3 patients that received TNK injection

Abbreviations: LKW, last known well; CT, computed tomography; TNK, tenecteplase; TD-NIRS, time-domain near-infrared spectroscopy

| Table S2. Median, Minimum, and Maximum Vitals During Thrombectomy | | | | |
|---|---|---|---|---|
| | **Pre-Recanalization (Median[IQR])** | **Post-Recanalization (Median [IQR])** | **Minimum % Change** | **Maximum % Change** |
| **MAP (mmHg)** | 91 (74-101) | 88 (72-98) | -0.68 % | -23% |
| **$SpO_2$ (%)** | 100 (97-100) | 100 (97-100) | 0.0% | 0.69 % |
| **$FiO_2$ (%)** | 56 (45-65) | 56 (45-64) | 0.0% | 15 % |
| **$ETCO_2$ (mmHg)** | 36 (34-37) | 34 (33-37) | -0.45% | -6.5% |

Abbreviations: MAP, mean arterial pressure; $SpO_2$, oxygen saturation; $FiO_2$, fraction of inspired oxygen; $ETCO_2$, end-tidal carbon dioxide; IQR, interquartile range

Median values with interquartile range, as well as the minimum and maximum percent change for the average MAP, $SpO_2$, $FiO_2$, and $ETCO_2$ 5 minutes before and after the recanalization time were determined. Out of 11 patients, A-line data was used for 8, while cuff data was used for the remaining 3 due to the unavailability or inaccuracy of their A-line data. Wilcoxon matched pairs signed-rank exact test revealed no significant differences in MAP ($r$=-0.32, $p$=0.31), $SpO_2$ ($r$=0.25, $p$=0.50), $FiO_2$ ($r$=0.15, $p$=0.68), and $ETCO_2$ ($r$=-0.34, , $p$=0.29) values pre- vs post-recanalization ($n$=11).

### Table S3. Perioperative Neuroimaging of All Patients Undergoing Thrombectomy

| Patient ID | Pre-EVT CTH | Occluded Vessel | Intra-EVT Angiography | | Post-EVT MRI | | Average $HbO_2$/HHb |
|---|---|---|---|---|---|---|---|
| Patient 1 | 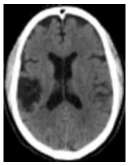 | L ICA | 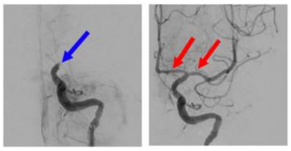 | EVT of the L ICA terminus occlusion (TICI 3). | 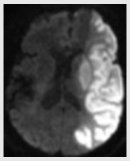 | L MCA territory infarct. | 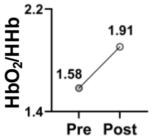 |
| Patient 2 | 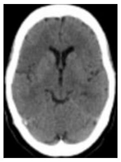 | R M2 | 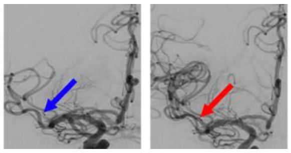 | EVT of the R superior division M2 branch MCA (TICI 3). | 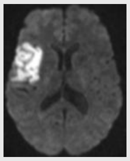 | R MCA territory acute infarct. | 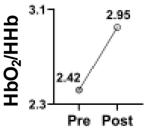 |
| Patient 3 | 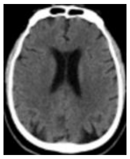 | R MCA and ICA Tandem | 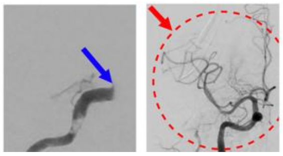 | EVT of the R ICA and the R MCA M1 segment (TICI 2B). | 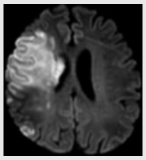 | Acute R MCA territory infarct with hemorrhagic transformation. | 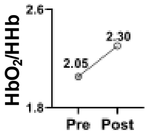 |
| Patient 4 | 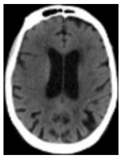 | R M2 | 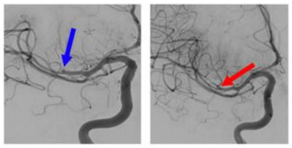 | EVT of the R MCA M2 (TICI 3). | 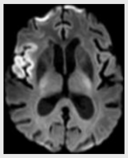 | Acute infarction of the R frontal lobe and insula. | 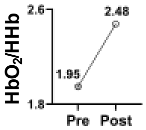 |
| Patient 5 | 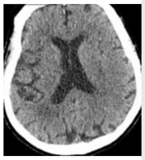 | R MCA and ICA Tandem | 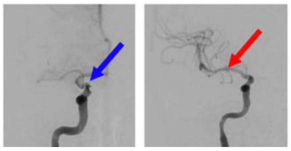 | EVT of the R ICA and MCA M1 (TICI 3) | 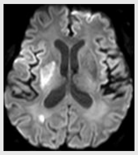 | R MCA territory infarct with petechial hemorrhage. | 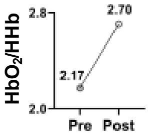 |
| Patient 6 | 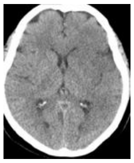 | L M1 | 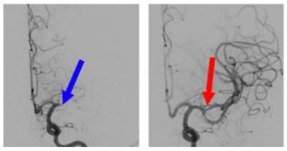 | EVT of L MCA M1. Stenosis at superior division (TICI 2C). | 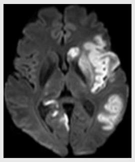 | Acute infarcts in the L MCA and R PCA territories. | 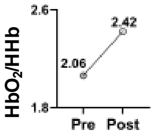 |
| Patient 7 | 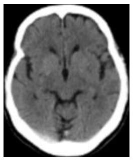 | L M1 | 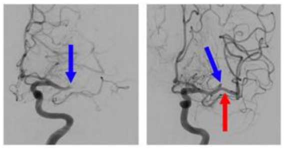 | EVT of the L M1 segment. Residual occlusion in L MCA M2 (TICI 2B). | 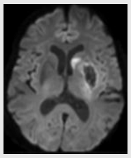 | L basal ganglia infarction with associated hemorrhagic conversion. | 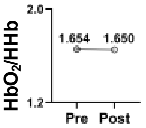 |
| Patient 8 | 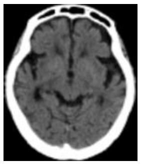 | R M2 | 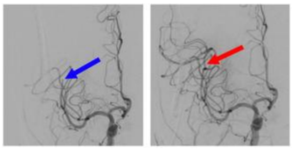 | EVT of the R MCA M2 (TICI 3). | 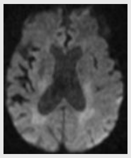 | No infarction or other acute intracranial abnormality. | 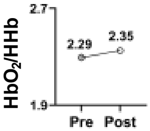 |
| Patient 9 | 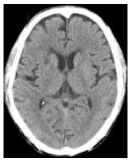 | R M1 | 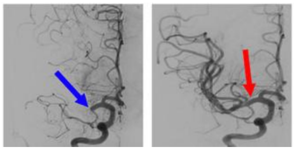 | EVT of the R MCA M1 (TICI 3). | 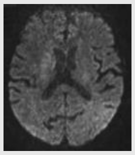 | Minimal R basal ganglia subacute-chronic infarction. | 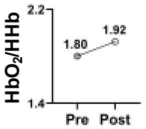 |
| Patient 10 | 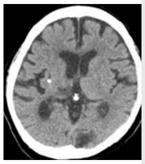 | L M1 | 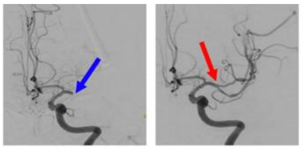 | EVT of the L MCA M1 (TICI 3). | 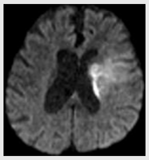 | Mild hemorrhagic conversion of the L basal ganglia infarct. | 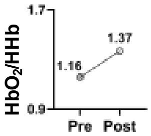 |

| Table S3. Perioperative Neuroimaging of All Patients Undergoing Thrombectomy (Continued) | | | | | | | |
|---|---|---|---|---|---|---|---|
| **Patient ID** | **Pre-EVT CTH** | **Occluded Vessel** | **Intra-EVT Angiography** | | **Post-EVT MRI** | | **Average $HbO_2/HHb$** |
| **Patient 11** | 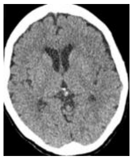 | L M1 | 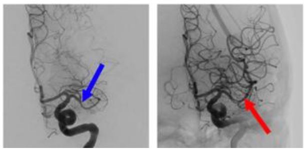 | EVT of the L MCA M1 (TICI 3). | 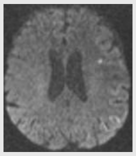 | Acute and punctate infarction in the L frontal lobe. | 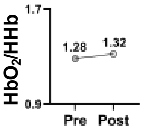 |

Abbreviations: CTH, CT Head; EVT, endovascular thrombectomy; MRI, magnetic resonance imaging; MCA, middle cerebral artery; ICA, internal carotid artery; [$HbO_2$], oxyhemoglobin concentration (μM); [HHb], deoxyhemoglobin concentration (μM).
Blue Arrow - occluded vessel
Red Arrow - open vessel

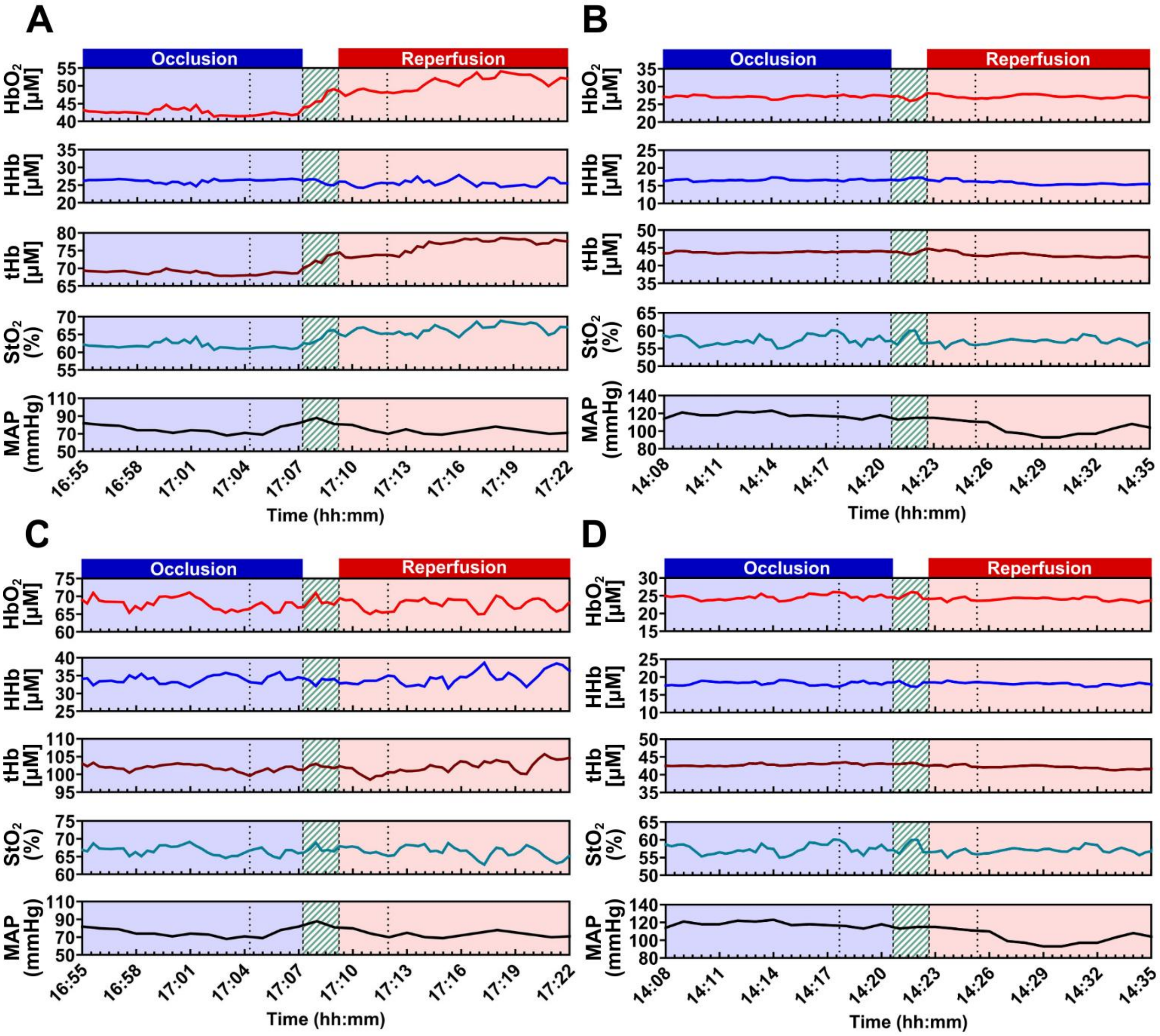


**Figure S1: Time series of TD-NIRS values for the full recording duration for two representative patients.** (A) The affected side for patient 1 and (B) patient 7 is represented including, [$HbO_2$], [HHb], [tHb], $StO_2$, and MAP. (C) The unaffected side for patient 1 and (D) patient 7 during EVT is shown. Dotted lines indicate the 2.5-minute pre- and post-recanalization data intervals used for analysis. Green shaded regions represent the two-minute recanalization window, centered at the recanalization time, which was excluded from analysis. [$HbO_2$], oxyhemoglobin concentration (μM); [HHb], deoxyhemoglobin concentration (μM); [tHb], total hemoglobin concentration (μM); $StO_2$, tissue oxygen saturation [%]; MAP, mean arterial pressure (mmHg); [μM] = micromolar.

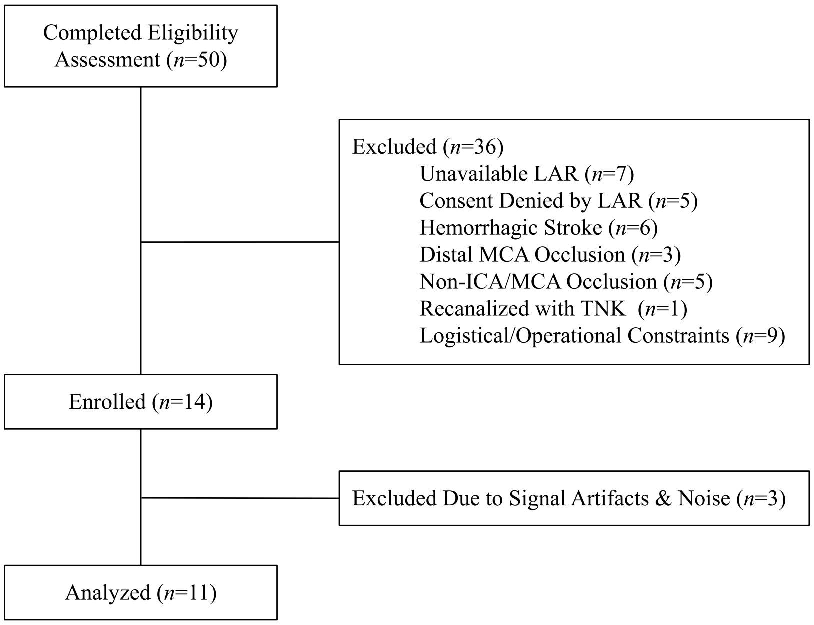


**Figure S2: Schematic representation of patient screening and enrollment.** LAR, legally authorized representative; MCA, middle cerebral artery; ICA, internal carotid artery; TNK, tenecteplase.
